# Chronic pain and differential brain morphology alterations of childhood maltreatment, trauma exposure in adulthood and their cumulative effects in the UK Biobank

**DOI:** 10.1101/2025.11.12.25339964

**Authors:** Tong-En Lim, William R. Reay, Sylvia M. Gustin, Yann Quidé

**Affiliations:** NeuroRecovery Research Hub, School of Psychology, The University of New South Wales (UNSW) Sydney, Sydney, NSW, Australia; Centre for Pain IMPACT, Neuroscience Research Australia (NeuRA), Randwick, NSW, Australia; Menzies Institute for Medical Research, University of Tasmania, Hobart, Tasmania, Australia

**Keywords:** Chronic pain, childhood maltreatment, brain morphology, trauma exposure, older adults, UK Biobank

## Abstract

**Background:** Chronic pain is associated with heightened prevalence of trauma exposure. Separate studies of chronic pain and trauma report overlapping brain alterations in older adults (age over 60). Despite this close relationship, the interplay between chronic pain and trauma on brain morphology remains poorly understood.

**Methods:** Tabulated T1-weighted structural magnetic resonance imaging data were accessed from the UK biobank (N = 21,996). Linear mixed models were used to determine the main effects of group (control versus chronic pain), severity of trauma exposure and their interaction on measures of surface area, cortical thickness and subcortical volume. To examine the unique and cumulative impacts of trauma exposure at different developmental timing, group of participants exposed to childhood maltreatment only, trauma in adulthood only and both were derived.

**Results:** Independently of the severity of trauma exposure, chronic pain was significantly associated with smaller surface area and thicker frontal, temporal, parietal, somatosensory, occipital and cingulate cortices. Independently of chronic pain, the severity of trauma exposure in adulthood and both childhood and adulthood, but not childhood alone, were associated with smaller bilateral nucleus accumbens, putamen, thalamus and left hippocampal volumes, as well as left inferior parietal gyrus surface area. No chronic pain-by-trauma severity interactions were significantly associated with variations in any regional measures.

**Conclusions:** Specific effects of chronic pain and trauma severity were evident among older adults exposed to trauma at different developmental timing. However, there was no evidence for unique brain signatures representing the combined effects of chronic pain and trauma in older adults.

## 1. Introduction

Chronic pain disproportionately affects older adults (age over 60) (1), with a large proportion (10% to 28%) of older adults with chronic pain reporting heightened lifetime levels of exposure to traumatic experiences (2). The prevalence of post-traumatic stress disorder (PTSD) in older adults varies between 10% and 50% across different chronic pain conditions, relative to approximately 3.9% in the general population (2,3). Consequently, this population of older adults usually experience worse outcomes including mobility limitations, decreased quality of life and cognitive decline (2).

Brain atrophy is typically observed in aging populations (4). When compared to typically aging adults, older adults with chronic pain (e.g., chronic back pain, headache/migraine, joint pain, craniomandibular disorder) show accelerated widespread brain atrophy including smaller cingulate, prefrontal and motor/premotor volumes, as well as smaller hippocampal volume that can exacerbate with aging (5,6). Accelerated reduction in grey matter volume of the orbitofrontal cortex was also found in “older” (40-49 years) compared to “younger” (20-29 years) adults with chronic pain (7), highlighting the critical need to understand underlying neural mechanisms of chronic pain in the aging population.

Interestingly, older adults with a history of trauma exposure show brain alterations similar to those observed in chronic pain, especially in stress-sensitive regions such as the frontal lobe, anterior cingulate cortex and subcortical regions including the hippocampus and amygdala (8–10). The timing of traumatic exposure can have differential impacts on brain morphology; for instance, amygdala volume is negatively associated with exposure to childhood maltreatment but positively associated with trauma exposure in adulthood in females with PTSD (11). Despite evidence for differential effects of trauma exposure on brain morphology at different developmental period, studies examining both childhood and adulthood trauma separately in the same sample remain limited.

The relationship between chronic pain and trauma exposure has been well established in older adults (2). Relative to controls, people with chronic pain show differential patterns of brain alterations depending on the severity of post-traumatic stress symptoms in core regions for trauma and pain processing, including the middle cingulate cortex, posterior insula and putamen (12). Despite sharing similar brain alterations, evidence on the combined effects of trauma and chronic pain on the brain remains scarce and limited to specific chronic pain populations (12).

The present study aims to determine the effects of trauma exposure at different neurodevelopmental stages, as well as their cumulative effects, on brain morphology in older adults from the general population who experience chronic pain. More pronounced grey matter reductions were hypothesised in older adults with chronic pain exposed to trauma, particularly in stress-sensitive regions (i.e., cingulate cortex, prefrontal cortex, insula, hippocampus). Furthermore, differential impacts of trauma depending on the developmental timing of exposure were also expected.

## 2. Methods and materials

### 2.1 Study population

This study accessed data from the UK Biobank (UKB; release v19) (13) to conduct secondary analyses (application ID 363586). The UKB is a very large, population-based prospective study that collected extensive phenotypic and genotypic information from over 500,000 British participants aged 40-69 when recruited in 2006-2010. Participants were excluded if they (1) did not participate in brain MRI scan at the first imaging visit (Instance 2), (2) did not respond/had incomplete responses to pain– and trauma-related questionnaires and/or (3) have a diagnosis of neoplasm, diseases of the nervous system, obesity, diabetes and/or narrow-band traumatic brain injury (International Statistical Classification of Diseases and Related Health Problems – version 10 [ICD-10] C00-D48, G00-G99, E65-E68, E10-E14, S00-S09). ICD-10–based exclusions were implemented to focus on pain-specific supraspinal mechanisms and to minimise confounding effects associated with potential brain injury or dysfunction.

### 2.2 Demographic information

Age at first imaging visit (field ID 21003), sex (field ID 31) and ethnicity (field ID 21000) were documented. Townsend deprivation index (field ID 22189) was administered at baseline to measure area-based social deprivation (accounting for unemployment, overcrowding, non-car ownership and non-home ownership), based on data from the preceding national census (14), where higher (more positive) score represents higher levels of deprivation. Diagnoses of mental and behavioural disorders (F00-F99) according to the ICD-10 criteria (field ID 41270) were recorded. Due to the large psychiatric comorbidities in chronic pain, we did not exclude participants reporting diagnoses of mental and behavioural disorders, but we accounted for their presence in focal analyses.

### 2.3 Chronic pain

Participants were asked about “pain types experienced in the last month” (field ID 6159) at the first imaging visit. The possible answers were pain at seven specific body sites (head, face, neck/shoulder, back, stomach/abdomen, hip, knee), pain all over the body, “none of the above” and “prefer not to answer”. Participant who answered “none of the above” were placed into the control (no pain) group and those who answered “prefer not to answer” were excluded from further analyses. Participants who indicated presence of pain at any site(s) were then asked if the pain had persisted for three or more months (category ID 100048). Participants who provided positive responses to having pain for three or more months were classified as having chronic pain, consistent with previous studies (15), whereas those who provided negative responses (i.e., experiencing acute pain) were excluded from further analyses.

### 2.4 Childhood maltreatment, adulthood and cumulative trauma

Childhood maltreatment was measured using the Childhood Trauma Screener (CTS-5) (16,17). The CTS-5 (field IDs 20487-91) consists of five items each describing physical, sexual and emotional abuse and physical and emotional neglect (for details, see Supplementary Table S1). Participants had to respond using a 5-point Likert scale: ‘0 = Never True’, ‘1 = Rarely True’, ‘2 = Sometimes True’, ‘3 = Often’, ‘4 = Very often true’. Two positively worded questions were reverse coded. The five items were summed to compute an overall childhood maltreatment severity score (range = 0-20; higher score represents higher severity). Severity of trauma exposure in adulthood (after the age of 16) was measured using an equivalent screener (field IDs 20521-25) adapted from a United Kingdom national crime survey for being a victim of crime and adult domestic violence (16,18). The screener consists of five questions describing relationship and financial insecurity, and physical, psychological and sexual intimate partner violence, measured using the same 5-point Likert scale: ‘0 = Never True’, ‘1 = Rarely True’, ‘2 = Sometimes True’, ‘3 = Often’, ‘4 = Very often true’ (for details see Supplementary Table S1). Two positively worded questions were reverse coded. Items reflecting the severity of interpersonal trauma exposure were summed to compute an overall adulthood trauma severity score (range = 0-16; higher score represents higher severity). The item reflecting financial insecurity was excluded from these analyses. Both severity scores were converted to z-scores and summed to compute an overall cumulative trauma severity score to ensure each questionnaire contributed similar weight. To examine the isolated impacts of childhood maltreatment, adulthood and cumulative trauma, participants who were exposed to childhood maltreatment only (i.e., not reporting being exposed to trauma in adulthood) were included in analyses of childhood maltreatment; participants who were exposed to adulthood trauma only (i.e., not reporting being exposed to childhood maltreatment) were included in analyses of adulthood trauma; those who were exposed to both childhood maltreatment and adulthood trauma were included in analyses of cumulative trauma. There was no overlap of participants among these trauma-related categories (i.e., all participants uniquely contributed to one statistical model).

### 2.5 MRI acquisition and processing

Tabulated T1-weighted data at first imaging visit (Instance 2) were accessed. Three-dimensional T1-weighted magnetisation-prepared rapid acquisition gradient echo scans were acquired using 3T Siemens Skyra scanners with a 32-channel head coil across four scanning sites using the following parameters: sagittal orientation; in-plane acceleration factor = 2; inversion time/repetition time = 880/2000 ms; voxel resolution = 1 × 1 × 1 mm; acquisition matrix = 208 × 256 × 256 mm (13). All neuroimaging data were minimally pre-processed with FreeSurfer (19) using protocols documented in Miller et al. (2016). Indices of regional subcortical volume (automated segmentation, *aseg*) (20), cortical thickness and surface area (Desikan-Killiany atlas) (21) were extracted.

### 2.6 Statistical analyses

All analyses were performed in R (v 4.4.0; R Core Team, 2023) on the UKB research analysis platform DNAnexus. Linear mixed models, using the “lme” function from the “nlme” package (v3.1-168; Pinheiro et al., 2025), were applied to analyse the imaging data. Group (chronic pain versus controls) and trauma severity scores (separately for childhood maltreatment, adulthood and cumulative trauma) were modelled as fixed effects. Subject ID and imaging sites were modelled as random effects. Age, squared mean-centred age (age^2^; to account for non-linear effects of age), sex, Townsend deprivation index and presence of mental health diagnoses were included as covariates. Total intracranial volume was included in analyses of cortical surface area and subcortical volume (but not cortical thickness), as these metrics are sensitive to brain size. Focal analyses estimated the main effects of group (chronic pain versus controls), trauma severity (separately for childhood, adulthood, cumulative) and their interactions (the product of group by mean-centred trauma severity score). False discovery rate (FDR) correction using Benjamini-Hochberg method (24) was applied to account for the number of regions-of-interest (ROIs) studied (66 ROIs for cortical thickness and surface area; 14 ROIs for subcortical volume). Statistical significance was set at *pFDR* < 0.05.

## 3. Results

### 3.1 Participant characteristics

Table 1 presents the demographic characteristics of the participants. Among the total sample (N = 21,996), 13,249 individuals (60.2%; 52.6% female) were assigned to the control group and 8,747 individuals (39.8%; 58.6% female) were assigned to the chronic pain group. Within the control group, 2,515 individuals (18.9%) reported being exposed to childhood maltreatment only, 2,553 (19.3%) exposed to adulthood trauma only, and 4,982 (37.6%) exposed to both. Within the chronic pain group, 1,609 individuals (18.4%) reported being exposed to childhood maltreatment only, 1,510 (17.3%) exposed to adulthood trauma only and 3,888 (44.5%) were exposed to both. The chronic pain group was significantly younger, reported significantly more severe childhood maltreatment, adulthood and cumulative trauma, included significantly more females and a higher proportion of participants with diagnoses of psychiatric comorbidities than the control group.

**Table 1.**
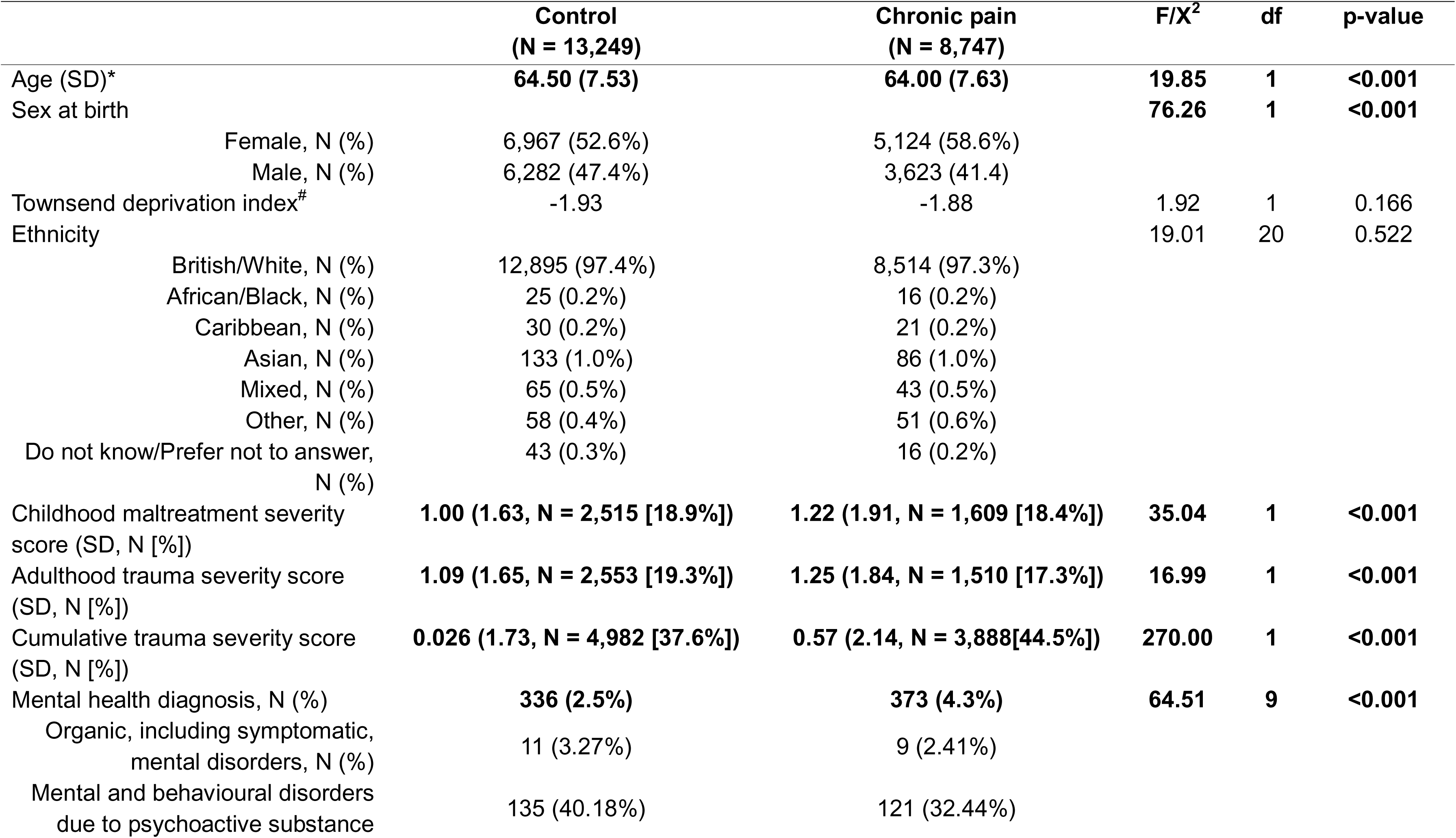
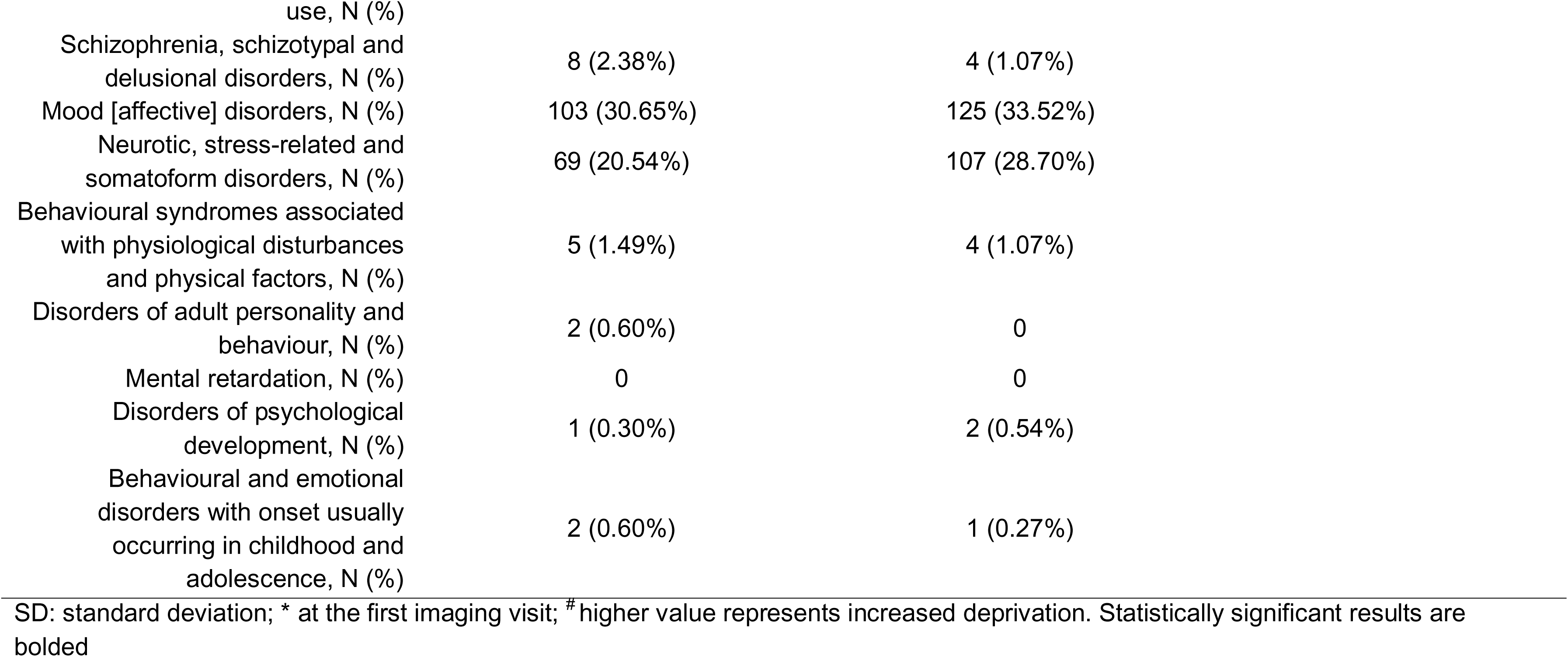
Demographic information.

| **Table 1. Demographic information** |  |  |  |  |  |
| --- | --- | --- | --- | --- | --- |
|  | **Control**<br>**(N = 13,249)** | **Chronic pain**<br>**(N = 8,747)** | **F/X<sup>2</sup>** | **df** | **p-value** |
| Age (SD)* | **64.50 (7.53)** | **64.00 (7.63)** | **19.85** | **1** | **<0.001** |
| Sex at birth |  |  | **76.26** | **1** | **<0.001** |
| Female, N (%) | 6,967 (52.6%) | 5,124 (58.6%) |  |  |  |
| Male, N (%) | 6,282 (47.4%) | 3,623 (41.4) |  |  |  |
| Townsend deprivation index<sup>#</sup> | -1.93 | -1.88 | 1.92 | 1 | 0.166 |
| Ethnicity |  |  | 19.01 | 20 | 0.522 |
| British/White, N (%) | 12,895 (97.4%) | 8,514 (97.3%) |  |  |  |
| African/Black, N (%) | 25 (0.2%) | 16 (0.2%) |  |  |  |
| Caribbean, N (%) | 30 (0.2%) | 21 (0.2%) |  |  |  |
| Asian, N (%) | 133 (1.0%) | 86 (1.0%) |  |  |  |
| Mixed, N (%) | 65 (0.5%) | 43 (0.5%) |  |  |  |
| Other, N (%) | 58 (0.4%) | 51 (0.6%) |  |  |  |
| Do not know/Prefer not to answer,<br>N (%) | 43 (0.3%) | 16 (0.2%) |  |  |  |
| Childhood maltreatment severity<br>score (SD, N [%]) | **1.00 (1.63, N = 2,515 [18.9%])** | **1.22 (1.91, N = 1,609 [18.4%])** | **35.04** | **1** | **<0.001** |
| Adulthood trauma severity score<br>(SD, N [%]) | **1.09 (1.65, N = 2,553 [19.3%])** | **1.25 (1.84, N = 1,510 [17.3%])** | **16.99** | **1** | **<0.001** |
| Cumulative trauma severity score<br>(SD, N [%]) | **0.026 (1.73, N = 4,982 [37.6%])** | **0.57 (2.14, N = 3,888[44.5%])** | **270.00** | **1** | **<0.001** |
| Mental health diagnosis, N (%) | **336 (2.5%)** | **373 (4.3%)** | **64.51** | **9** | **<0.001** |
| Organic, including symptomatic,<br>mental disorders, N (%) | 11 (3.27%) | 9 (2.41%) |  |  |  |
| Mental and behavioural disorders<br>due to psychoactive substance | 135 (40.18%) | 121 (32.44%) |  |  |  |

|  |  |  |
| --- | --- | --- |
| use, N (%) |  |  |
| Schizophrenia, schizotypal and delusional disorders, N (%) | 8 (2.38%) | 4 (1.07%) |
| Mood [affective] disorders, N (%) | 103 (30.65%) | 125 (33.52%) |
| Neurotic, stress-related and somatoform disorders, N (%) | 69 (20.54%) | 107 (28.70%) |
| Behavioural syndromes associated with physiological disturbances and physical factors, N (%) | 5 (1.49%) | 4 (1.07%) |
| Disorders of adult personality and behaviour, N (%) | 2 (0.60%) | 0 |
| Mental retardation, N (%) | 0 | 0 |
| Disorders of psychological development, N (%) | 1 (0.30%) | 2 (0.54%) |
| Behavioural and emotional disorders with onset usually occurring in childhood and adolescence, N (%) | 2 (0.60%) | 1 (0.27%) |

SD: standard deviation; \* at the first imaging visit; # higher value represents increased deprivation. Statistically significant results are bolded

### 3.2 Childhood maltreatment

Statistical details for the analyses investigating the effects of group, severity of childhood maltreatment and their interactions are reported in Supplementary Tables S2-S4. When accounting for the severity of childhood maltreatment exposure only, chronic pain was significantly associated with smaller surface area in the bilateral fusiform and postcentral gyri, in the left bank of the superior temporal sulcus and posterior cingulate gyrus, and in the right insula, middle temporal, superior frontal and superior parietal gyri (see Figure 1). Chronic pain was also significantly associated with thicker right superior parietal gyrus, but not with variations in volumes for any of the subcortical ROIs. The severity of childhood maltreatment and group-by-maltreatment interaction were not significantly associated with variations in any of the ROIs studied.

**Figure 1.**
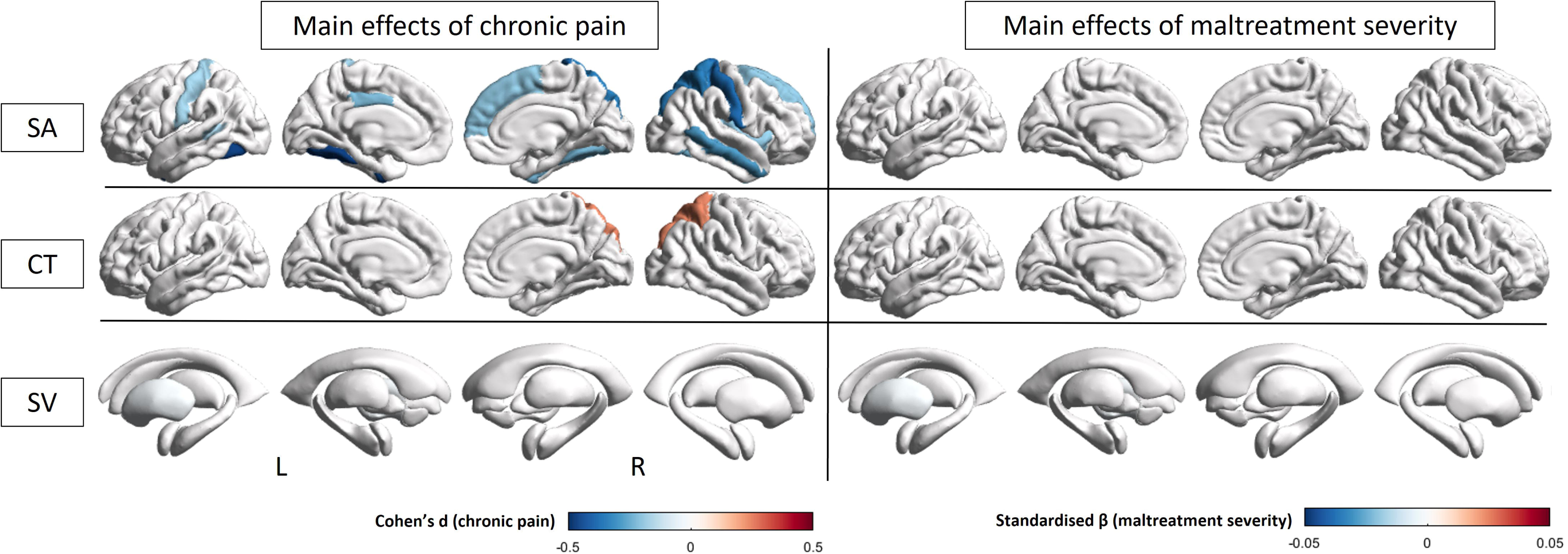
Significant effects of chronic pain and severity of childhood maltreatment exposure on regional grey matter measures. Significant differences in surface area (SA), cortical thickness (CT) and subcortical volume (SV) associated with chronic pain (left panel) and severity of childhood maltreatment exposure (right panel). Effect sizes are presented in standardised beta (trauma severity) and Cohen’s d (group); positive effect size (red) represents increase/positive relationship whereas negative effect size (blue) represents decrease/negative relationship.

### 3.3 Adulthood trauma

Statistical details for the analyses investigating the effects of group, severity of adulthood trauma exposure and their interactions are reported in Supplementary Tables S5-S7. When accounting for the severity of trauma exposure in adulthood only, chronic pain was significantly associated with smaller surface area in 18 ROIs, including the bilateral fusiform, middle and superior temporal, postcentral gyri, as well as the left inferior parietal, inferior temporal, lateral occipital gyri and right caudal and rostral anterior cingulate, posterior cingulate, insula, superior frontal, superior parietal and supramarginal gyri. In addition, chronic pain was significantly associated with thicker bilateral caudal and rostral middle frontal, superior frontal, inferior and superior parietal, lingual, pars triangularis, as well as left lateral occipital, and right precentral, precuneus and supramarginal gyri (see Figure 2). Chronic pain was not significantly associated with variations in volume for any of the subcortical ROIs. The severity of adulthood trauma exposure was significantly associated with smaller bilateral nucleus accumbens, putamen and thalamus, and left hippocampal volumes. The group-by-trauma interaction was not significantly associated with variations in any of the ROIs studied.

**Figure 2.**
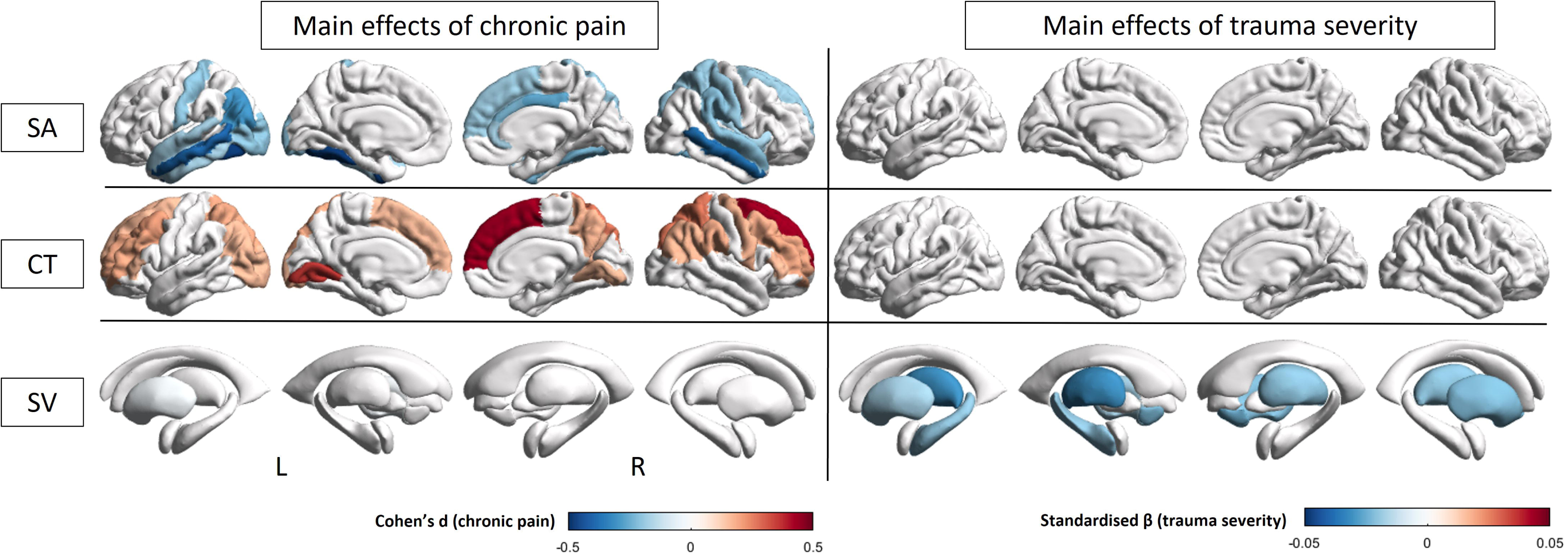
Significant effects of chronic pain and severity of trauma exposure in adulthood on regional grey matter measures. Significant differences in surface area (SA), cortical thickness (CT) and subcortical volume (SV) associated with chronic pain (left panel) and severity of trauma exposure in adulthood (right panel). Effect sizes are presented in standardised beta (trauma severity) and Cohen’s d (group); positive effect size (red) represents increase/positive relationship whereas negative effect size (blue) represents decrease/negative relationship.

### 3.4 Cumulative trauma

Statistical details for the analyses investigating the effects of group, severity of cumulative trauma exposure and their interactions are reported in Supplementary Tables S8-S10. When accounting for the severity of exposure to both childhood maltreatment and trauma in adulthood, chronic pain was significantly associated with smaller surface area in 27 ROIs, including the bilateral fusiform, inferior and middle temporal, lingual, pericalcarine, postcentral, posterior cingulate, precentral, superior frontal gyri, as well as the left bank of the superior temporal sulcus, inferior parietal, lateral occipital, right caudal and rostral anterior cingulate, insula, pars triangularis, superior parietal and superior temporal gyri (see Figure 3). In addition, chronic pain was significantly associated with thicker bilateral lateral occipital, pericalcarine, precuneus, superior parietal, and right rostral middle frontal gyri. The severity of cumulative trauma was significantly associated with smaller left inferior parietal surface area and left putamen volume. The group-by-trauma interaction was not significantly associated with variations in any of the ROIs studied.

**Figure 3.**
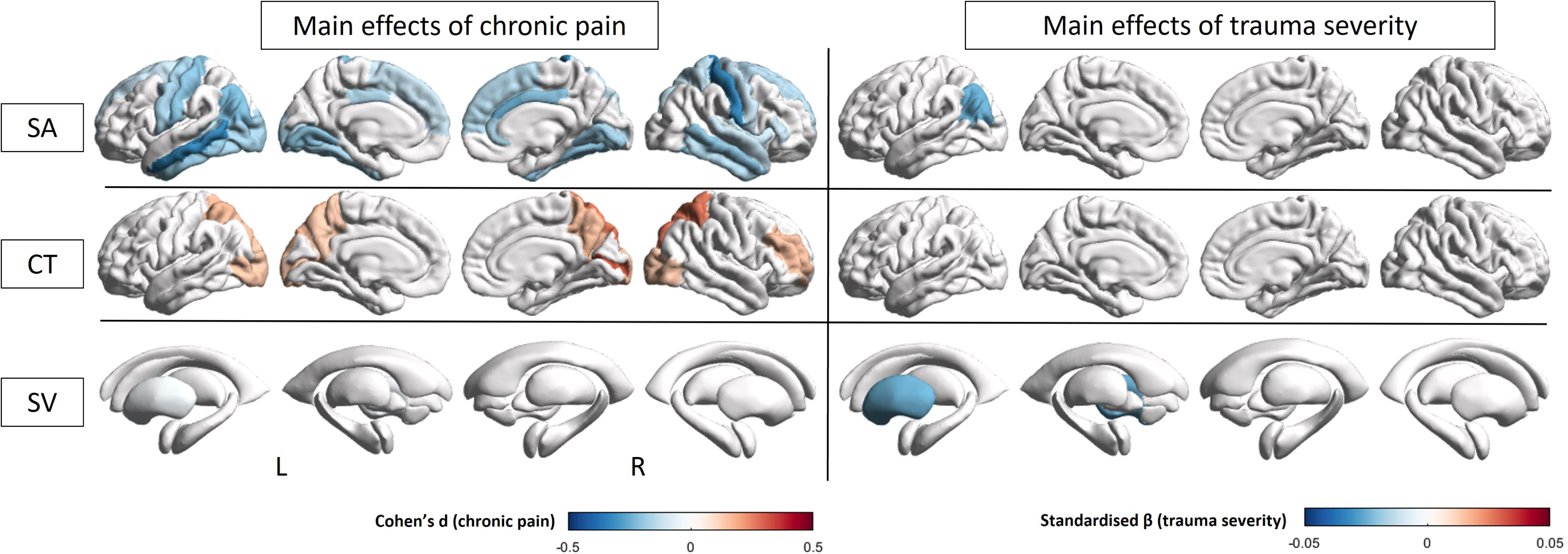
Significant effects of chronic pain and cumulative severity of childhood maltreatment and trauma exposure in adulthood on regional grey matter measures. Significant differences in surface area (SA), cortical thickness (CT) and subcortical volume (SV) associated with chronic pain (left panel) and cumulative severity of childhood maltreatment and trauma exposure in adulthood (right panel). Effect sizes are presented in standardised beta (trauma severity) and Cohen’s d (group); positive effect size (red) represents increase/positive relationship whereas negative effect size (blue) represents decrease/negative relationship.

## Discussion

The present study examined the effects of chronic pain, the severity of trauma exposure as well as their interactive effects on brain morphology in older adults. Consistent with previous studies (15,25), chronic pain was associated with smaller surface area and thicker superior frontal, temporal, superior parietal, somatosensory, occipital, cingulate and insular cortices. On the other hand, independently of chronic pain, the severity of childhood maltreatment was not associated with variations in any ROIs studied. The severity of trauma exposure in adulthood was associated with smaller nucleus accumbens, putamen, thalamus and hippocampal volumes. The cumulative severity of childhood maltreatment and trauma exposure in adulthood was associated with smaller left inferior parietal gyrus surface area and putamen volume. No group-by-trauma severity interaction (separately for childhood maltreatment, adulthood trauma or both) was associated with variations in any ROIs studied, indicating that the severity of trauma exposure, independent of timing or accumulation of exposure, did not differentially impact the brains of older adults with chronic pain.

Partially consistent with our hypotheses, childhood maltreatment, trauma exposure in adulthood and their cumulative effects had different impacts on brain morphology, particularly in the nucleus accumbens, putamen, thalamus, hippocampus and inferior parietal gyrus, independently of chronic pain. The subcortical regions are part of the prefrontal-limbic-striatal circuit, the core system implicated in trauma exposure and PTSD (26–28). The long-term expression of negative emotions, such as fear and anxiety, following more severe trauma exposure, may lead to neurobiological changes including reduced neurogenesis and hyperactivation within the prefrontal-limbic-striatal circuit (28). Trauma-related morphological and functional alterations can influence each other, potentially reinforcing maladaptive stress-related neural responses (29,30). Smaller volume of these subcortical regions has commonly been associated with heightened stimulus salience/sensitivity (i.e., reward and emotion properties) and altered affective states following traumatic experiences in PTSD (28), potentially contributing to emotion dysregulation symptoms in individuals exposed to trauma (31,32). Interestingly, severity of trauma exposure in adulthood only, was associated with more pronounced volumetric reduction than cumulative trauma, when compared to controls. This may point to mechanisms of subsequent resilience to stressful/traumatic experiences, likely in terms of (blunted) stress reactivity and (improved) emotion regulation, following childhood maltreatment (33,34). Smaller inferior parietal surface area was uniquely associated with the severity of cumulative trauma and may represent a neural signature of repeated stress exposure. The inferior parietal gyrus, involved in spatial memory and visual-spatial processing, matures relatively later in development (35) Due to heightened levels of neuroplasticity as a result of a longer maturation period, the inferior parietal gyrus may be more vulnerable to chronic or repeated stress and subsequently, to later stress in adulthood (33,35). This suggests that morphological resilience may not be uniform across the brain; emotion-related regions may be more resilient following childhood maltreatment whereas higher-order regions (i.e., inferior parietal gyrus) can accumulate stress burden over the lifespan (36) This interpretation is consistent with differences in emotion dysregulation symptoms, depending on the timing of trauma exposure (37). Future longitudinal studies are needed to test this speculation.

These results are partially inconsistent with previous studies of trauma exposure in the UKB cohort. A previous study reported smaller cortical surface area in frontal and parietal regions as well as in the ventral diencephalon, hippocampus, thalamus and nucleus accumbens volumes in relation to childhood maltreatment exposure (10). Another study reported reduced cerebellar and ventral striatum volumes in participants exposed to childhood emotional abuse, or no significant brain alterations in relation to exposure to trauma in adulthood (38). This discrepancy relative to our findings may, at least in part, be attributable to the fact that these previous studies did not account for trauma exposure during other developmental windows than the one studied. Our results provide some evidence supporting this speculation. First, the severity of trauma exposure in adulthood alone and the cumulative effects with childhood maltreatment were associated with similar variation in brain morphology to those presented in these previous studies (10,38). Consistent with Madden et al. (2023), results from the present study show that the severity of cumulative trauma was associated with smaller surface area in the left inferior parietal gyrus whereas the severity of trauma exposure in adulthood was associated with smaller hippocampus, thalamus and nucleus accumbens volumes, similar to the results reported by these previous studies in relation to childhood maltreatment (10,38). Second, these previous studies might have labelled participants exposed to both childhood maltreatment and trauma in adulthood, as exposed to childhood maltreatment or exposure to adulthood trauma, depending on their research question. This is evident in the number of participants exposed to cumulative trauma being higher than those exposed to childhood maltreatment or adulthood trauma alone in the present study. Importantly, the present study indicates that isolated impacts of adulthood and cumulative trauma on the brain may be more pronounced than those of childhood maltreatment alone in older adults when accounting for chronic pain. This highlights the importance of considering and accounting for lifetime trauma exposure when examining the relationship between brain morphology and trauma in this population.

In addition, the effects of chronic pain observed here were comparable to a previous study using a larger sample of the UKB cohort (15), but not accounting for trauma exposure. Chronic pain was overall associated with smaller surface area in the temporal, motor, somatosensory, occipital and superior parietal and frontal regions and thicker frontal, occipital, parietal regions. However, when accounting for childhood maltreatment, chronic pain was associated with fewer brain regions and showed smaller effect sizes compared to models accounting for adulthood and cumulative trauma. These variations introduced by the type of trauma accounted for in each model indicate that childhood maltreatment and chronic pain potentially have large overlapping effects on brain morphology. This overlap also highlight that different mechanisms are at play following exposure to adverse events in childhood or adulthood, involving stress perception, reactivity and regulation (39). Together, these results indicate that trauma/adversity exposure during different developmental windows have different impacts on brain morphology in the context of chronic pain. Finally, contrary to our hypotheses, no group-by-trauma interactions were significantly associated with variations in brain morphology, indicating that the associations between chronic pain and brain morphology did not depend on the severity of trauma experienced in this population.

This study has several limitations. First, the distribution of trauma severity scores showed a limited range and was heavily skewed towards lower values (see Supplementary Figure S1), indicating restricted variability in this population. Second, the exact timing of trauma exposure could not be pinpointed due to the types of questionnaires administered. In the UKB cohort, childhood maltreatment was measured using the CTS-5, a very short version of the Childhood Trauma Questionnaire (40), for exposure any time before the age of 16 whereas the severity of trauma exposure in adulthood was assessed using an equivalent screener adapted from a UK national crime survey, without providing an estimate of timing other than exposure after the age of 16. Accounting for exposure at specific developmental timing is critical as recent and long-term traumatic exposure can have different impacts on the brain (41). In addition, these tools are self-report questionnaires that may be subject to recall bias; however, previous studies suggested that self-report indices of trauma exposure demonstrate good reliability, even in samples of older adults with severe psychiatric conditions (42,43). Finally, participants in the UKB study were recruited from the general population and do not represent samples of medically assessed chronic pain participants. These limitations may partly explain the lack of group-by-trauma severity interactions in this population. Future studies of clinical populations that collect information on the exact timing of trauma exposure, especially during critical developmental periods in childhood and adolescence (44), are needed to better understand the role of timing of trauma exposure on brain morphology in this population.

Chronic pain and the severity of exposure to childhood maltreatment and/or trauma later in adulthood have differential impacts on brain morphology in older adults from the general population. Brain alterations associated with chronic pain were not dependent on the severity of trauma exposure in this population. Although these findings indicate that chronic pain and trauma may not have additive effects in older adult populations, it highlights the importance of incorporating trauma-informed care, regardless of chronic pain status, to better improve outcomes.

## Acknowledgements

This research has been conducted using the UKB resource (application no. 363586). Tong-En Lim was supported by a University of New South Wales (UNSW Sydney) International Postgraduate Scholarship and Edward C. Dunn scholarship administered by Neuroscience Research Australia (NeuRA). William R. Reay was supported by an NHMRC Emerging Leadership Investigator Grant. Sylvia M. Gustin was supported by Rebecca Cooper Fellowship from the Rebecca L. Cooper Medical Research Foundation. We are grateful to the study participants for their time and participation.

## Data availability

The data from the UKB used in this study are publicly available via their standard data access procedure at https://www.ukbiobank.ac.uk/.

## Authors’ contribution

Tong-En Lim contributed conceptualisation, data curation, formal analysis, methodology, visualization, validation, writing of the original draft, review and editing. William R. Reay contributed conceptualisation, methodology, and review and editing. Sylvia M. Gustin contributed conceptualisation, funding acquisition, methodology, project administration, resources, supervision, and review and editing. Yann Quidé contributed conceptualisation, data curation, formal analysis, methodology, visualisation, supervision, validation, writing of the original draft, review and editing.

